# Social isolation as a sex-specific risk factor for skeletal health in the Framingham Heart Study

**DOI:** 10.64898/2026.11.13.26368021

**Authors:** Scott Eugley, John DiPalazzo, P. Emogene Miller, Brooklynn M. Merrill, Eleanor Buchanan, Kristin Svahn, Christine W. Lary, Rebecca V. Mountain

## Abstract

**Importance:** Social isolation (SI) is a highly prevalent modifiable risk factor for numerous diseases and has been linked to worsened skeletal health. However, evidence from preclinical models suggests the effects of SI on skeletal health may be sex-specific.

**Objective:** To investigate if objective and subjective SI are associated with skeletal health differentially among older adult women and men.

**Design:** This prospective cohort study used SI and bone health data from the Framingham Heart Study Offspring Cohort, a community-based cohort, collected from 1998 to 2014. Analyses were conducted from 2023-2026.

**Participants:** 2,850 adult women and men were included in this study.

**Exposures:** Objective SI was measured using the Berkman-Syme Social Network Index, and subjective SI (i.e., loneliness) was measured using the Center for Epidemiological Studies Depression Scale “Loneliness” subscale.

**Main Outcomes and Measures:** Fragility fractures, bone mineral density, and bone turnover markers were assessed from clinic data.

**Results:** This study included 2,850 adults (median [IQR] age, 60 [53-67], 1,553 [54%] female; 2,728 [99%] white). Objective SI and loneliness were more prevalent among women. Loneliness was associated with increased risk for fracture (OR: 2.46, 95% CI: 1.25-4.57, *P* = 0.011) and decreased tibial bone mineral density (β = -14.13, 95% CI: -28.03 to -0.23, *P =* 0.046) in men, but not women. Objective SI was associated with decreased total osteocalcin (β = -0.43, 95% CI: -0.75 to -0.10, *P =* 0.010), a marker of bone formation, in men, but associated with an increase in osteoprotegerin, another marker of bone formation, in women (β = 0.18, 95% CI: 0.01-0.34, *P =* 0.035). SI was not associated with differences in sex hormones, but objective SI was associated with higher 8-isoprostane in women (β = 0.177, 95% CI: 0.062-0.292, *P* = 0.003, *q* = 0.025) and IL-6 in men (β = 0.195, 95% CI: 0.085-0.304, *P =* 0.001, *q* = 0.005). There was no mediating effect of IL-6 on loneliness and male bone outcomes.

**Conclusions and Relevance:** Our findings recapitulate sex differences previously seen in preclinical models of SI and indicate SI is an important modifiable risk factor for skeletal health among older adult men.

**KEY POINTS:** *Question:* Are objective social isolation and loneliness negatively associated with skeletal health disproportionately in men compared to women?

*Findings:* In this cohort study of 2,850 adults, loneliness was associated with a significant increase in fracture risk and lower bone mineral density in men. Objective isolation was associated with lower bone formation markers in men, but higher bone formation markers in women.

*Meaning:* Social isolation may be an important modifiable risk factor for male skeletal health.

## Introduction

In 2023, the U.S. Surgeon General announced concern about the rising “epidemic of loneliness and isolation,” warning against the public health dangers of social isolation (SI)^1^. SI is highly prevalent, particularly among older adults, affecting 1 in 4 individuals above the age of 65 in the U.S.^2^. From 2003-2020, the average time Americans spent alone increased by 24 hours per month^3^, and the global prevalence of SI increased by over 13% since the onset of the COVID-19 pandemic^4^. SI can be divided into *objective* SI, characterized by a lack of social connections, and *subjective* SI, characterized by the feeling of loneliness. The negative effects of both objective SI and loneliness are substantial, increasing risk for mental health disorders^5^, dementia^6,7^, cardiovascular disease^8^, and metabolic disorders^9^. SI is also associated with a 30% increased risk in mortality^10^, comparable to chronic smoking^1^.

Several studies, including our own^11,12^, demonstrate that SI is similarly harmful to skeletal health in preclinical models^13–17^ and human cohorts^18–22^. In mice, SI worsens trabecular and cortical bone microarchitecture in males^11^. These effects occur rapidly, persist with long-term SI^23^, and cannot be explained by thermal stress from single housing^12^. Other preclinical studies have found that SI decreases bone mineral content and bone mineral density (BMD)^15^, decreases adaptive response to artificial loading^17^, and alters markers of osteoclast activity^13^. Comparable results have emerged from human studies, showing that SI is associated with lower BMD^21^, increased risk of osteoporosis^20–22,24^, osteopenia^18,21^, and fracture^19,22^. Furthermore, isolated individuals have longer hospital stays^25^ and worse outcomes^26,27^ *after* fracture, potentially establishing a vicious cycle of compromised bone health. These findings are particularly concerning for older adults who are already the most vulnerable to SI^2^ and skeletal disorders^28^.

While prior research has demonstrated SI as a risk factor for skeletal health, this effect may be sex-specific. In preclinical models, ourselves^11,23^ and others^17^ have found that SI worsens bone parameters in male but not female mice. However, whether the same is true in humans is unclear, as only two clinical studies^19,20^ have considered sex differences in SI and bone. Researchers found a significant interaction with sex in the effect of SI on BMD, with a worse effect for men, although the overall effect of SI on BMD was not significant^19^, and found stronger associations between SI and osteoporosis among men^20^. These findings are particularly notable as women typically report higher levels of SI and loneliness^29,30^, and are more vulnerable to osteoporosis and related fractures than men^28^, but clinical and preclinical studies suggest SI has worse health effects in males. SI has worse effects on brain health^31,32^, inflammation^32,33^, HPA-function^34^, and higher mortality risk in males^33^. Furthermore, older men are less likely to report feelings of loneliness than women^30^, which could mask SI as an important factor for men’s musculoskeletal health.

The mechanisms by which SI impacts skeletal health, and if sex hormones drive differences between men and women, are unknown. However, a recent study^22^ reported that inflammatory markers partially mediate the association between SI, osteoporosis, and fracture risk, although these effects accounted for less than 9% of any association between SI and bone, and they did not evaluate sex differences. Chronic inflammation may also mediate the worsened SI-induced effects on cognition^32^ and mortality^33^ in men versus women, demonstrating a potential role of inflammation in sex-dependent SI-induced skeletal changes.

The aim of the present study was to investigate how the effects of objective SI and loneliness on skeletal health may differ between women and men in a cohort of older adults from the Framingham Heart Study (FHS). We also investigated if the sex-dependent effects of SI could be explained by differences in sex hormones or inflammatory factors. To our knowledge, this is the first study to characterize the sex-specific association between SI and skeletal health outcomes in humans.

## Methods

### Study Sample

We analyzed data from the Framingham Heart Study (FHS) Offspring Cohort, a prospective cohort of older adults from Framingham, Massachusetts, specifically those members who were enrolled in the ancillary Framingham Osteoporosis Study^35^. All data were acquired via the NIHLBI’s Biologic Specimen and Data Repository Information Coordinating Center (Acc.No.HLB00060025a). Ethical approval was obtained from the MaineHealth Institutional Review Board. The Offspring Cohort began in 1971 and enrolled children of participants from the original FHS cohort. Our analyses used clinic and survey data collected from Exam 7 (1998-2001) through Exam 9 (2011-2014). Sex was recorded at Exam 1 as Female or Male. We use the terms female/woman/women and male/man/men interchangeably in this paper.

### Objective and Subjective Social Isolation Measures

Exposure to objective SI was evaluated using the Berkman–Syme Social Network Index (SNI)^36^ measured at Exam 7 treated as a binary variable (**eMethods**). Scores of 0-2 were categorized as “isolated” and scores of 3-4 were categorized as “not isolated.”^37^ Exposure to subjective SI (i.e., loneliness) was evaluated using the “Loneliness” measure in the Center for Epidemiological Studies Depression Scale (CES-D)^38^ measured at Exam 7 (**eMethods**). We treated this as a binary variable, with scores of 1-3 categorized as “lonely”, and 0 categorized as “not lonely.”^39,40^

### Bone Measures

Fragility fractures were assessed using self-reported fractures after Exam 7 (**eMethods**).

Volumetric bone mineral density (vBMD) of the radius and tibia was measured in mg HA/cm^3^ using peripheral quantitative computed tomography (pQCT) at Exam 9. Areal bone mineral density (aBMD) of the femoral neck and lumbar spine was measured in g/cm^2^ using dual energy x-ray absorptiometry (DXA) at Exam 8. Circulating bone turnover markers RANKL, osteoprotegerin (OPG), and total and uncarboxylated osteocalcin were measured in blood samples collected at Exams 7 and 8.

### Circulating Sex Hormones and Inflammatory Markers

Circulating sex hormones were measured via liquid chromatography-mass spectrometry in serum samples collected at Exam 7. Estradiol (pg/mL) was measured only in women. Free (pg/mL) and total (ng/dL) testosterone was measured in both sexes. Circulating inflammatory markers were measured via enzyme-linked immunosorbent assays in serum or plasma from fasting blood samples, or urine samples collected at Exam 8.

### Statistical Modeling

Univariate participant characteristics and demographics were summarized and stratified by sex. Continuous variables were summarized using medians and interquartile ranges, and categorical variables using counts and percentages. Binary fracture outcomes were modeled using Firth penalized logistic regression to reduce small-sample bias and address potential model separation in the sex-stratified models. Associations for fracture outcomes are reported as odds ratios with 95% confidence intervals.

Continuous outcomes, including aBMD and vBMD measures, bone turnover markers, sex hormones, and inflammatory markers were modeled using linear regression and are reported as beta coefficients with 95% confidence intervals. “Not isolated” and “Not lonely” individuals were used as the reference groups for objective SI and loneliness, respectively.

Model 1 was specified as the primary inferential model and adjusted for age and body mass index (BMI), given their strong relationships with skeletal health^41^. Model 0 was unadjusted and presented alongside Model 1 to assess the influence of age and BMI adjustment. For each sex, exposure, and outcome combination, Models 0 and 1 were estimated using the same fixed complete-case analytical sample, defined by non-missing exposure, outcome, age, and BMI data. A fully adjusted Model 2 was evaluated as a sensitivity analysis, including additional social and physiological covariates (**eMethods**). Additionally, we performed a causal mediation analysis to determine if inflammatory markers mediated the associations between SI and bone outcomes (**eMethods**).

Missing data were handled using complete-case analysis. All analyses were conducted in R, version 4.2.1 (2022-06-23). Statistical tests were two-sided, and p-values less than 0.05 were considered statistically significant.

## Results

### Sample Characteristics

2,850 adults (54% female, 99% White) from the FHS Offspring Cohort were included in our analyses, with an average age of 60 years (**Table 1**). 95.8% of the women in the sample were post-menopausal. Approximately half (51%) of the individuals were objectively isolated, while 17% were subjectively isolated (i.e., lonely) (**Table 2**). SI differed between sexes, with more women being both objectively isolated and lonely. Fragility fractures were likewise more common among women (11%) than men (4.5%).

**Table 1.**
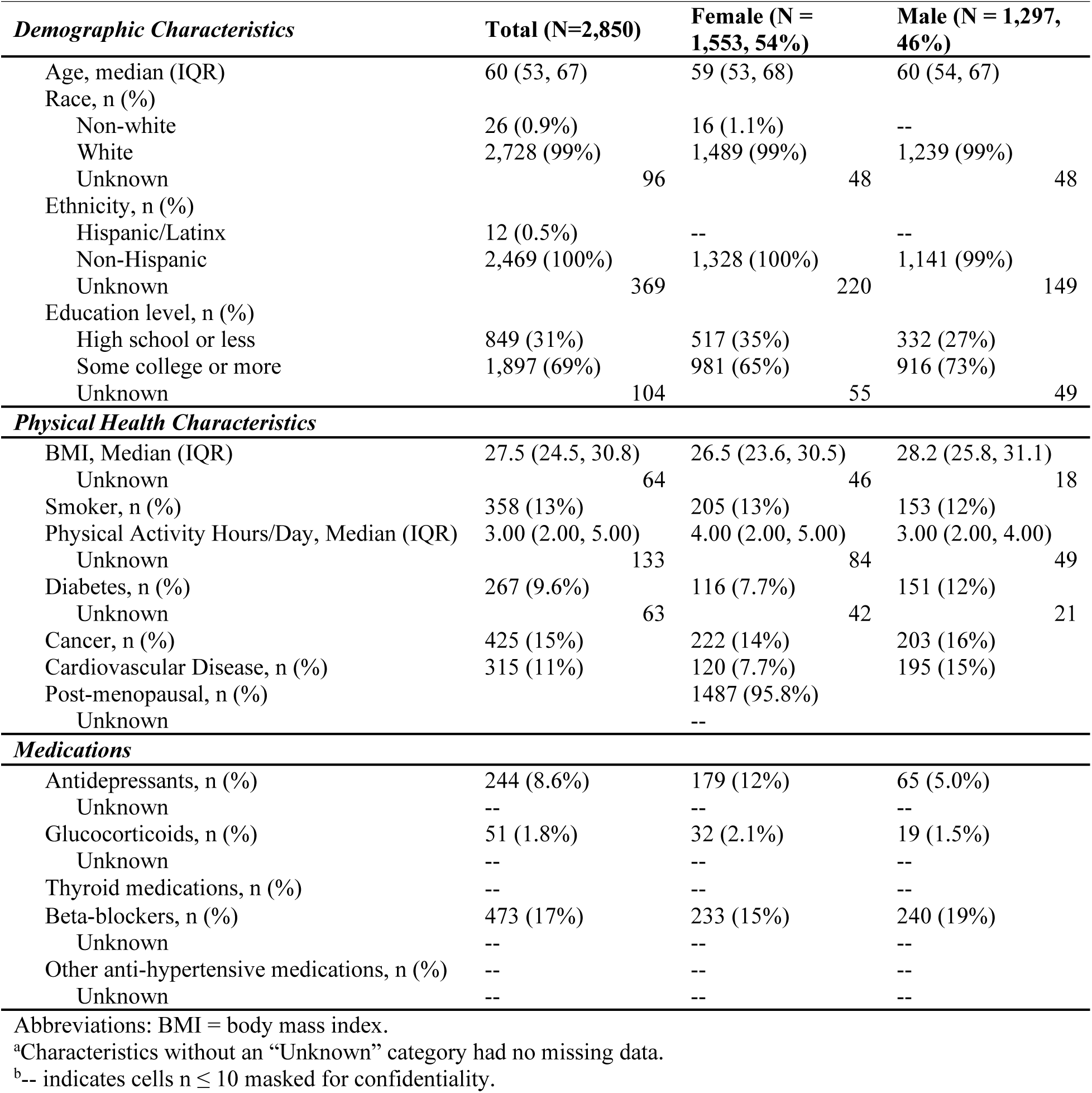
Demographic and health characteristics of the FHS analytic sample^a,b^.

| *Demographic Characteristics* | **Total (N=2,850)** | **Female (N = 1,553, 54%)** | **Male (N = 1,297, 46%)** |
|---|---|---|---|
| Age, median (IQR) | 60 (53, 67) | 59 (53, 68) | 60 (54, 67) |
| Race, n (%) |  |  |  |
| Non-white | 26 (0.9%) | 16 (1.1%) | -- |
| White | 2,728 (99%) | 1,489 (99%) | 1,239 (99%) |
| Unknown | 96 | 48 | 48 |
| Ethnicity, n (%) |  |  |  |
| Hispanic/Latinx | 12 (0.5%) | -- | -- |
| Non-Hispanic | 2,469 (100%) | 1,328 (100%) | 1,141 (99%) |
| Unknown | 369 | 220 | 149 |
| Education level, n (%) |  |  |  |
| High school or less | 849 (31%) | 517 (35%) | 332 (27%) |
| Some college or more | 1,897 (69%) | 981 (65%) | 916 (73%) |
| Unknown | 104 | 55 | 49 |
| ***Physical Health Characteristics*** |  |  |  |
| BMI, Median (IQR) | 27.5 (24.5, 30.8) | 26.5 (23.6, 30.5) | 28.2 (25.8, 31.1) |
| Unknown | 64 | 46 | 18 |
| Smoker, n (%) | 358 (13%) | 205 (13%) | 153 (12%) |
| Physical Activity Hours/Day, Median (IQR) | 3.00 (2.00, 5.00) | 4.00 (2.00, 5.00) | 3.00 (2.00, 4.00) |
| Unknown | 133 | 84 | 49 |
| Diabetes, n (%) | 267 (9.6%) | 116 (7.7%) | 151 (12%) |
| Unknown | 63 | 42 | 21 |
| Cancer, n (%) | 425 (15%) | 222 (14%) | 203 (16%) |
| Cardiovascular Disease, n (%) | 315 (11%) | 120 (7.7%) | 195 (15%) |
| Post-menopausal, n (%) |  | 1487 (95.8%) |  |
| Unknown |  | -- |  |
| ***Medications*** |  |  |  |
| Antidepressants, n (%) | 244 (8.6%) | 179 (12%) | 65 (5.0%) |
| Unknown | -- | -- | -- |
| Glucocorticoids, n (%) | 51 (1.8%) | 32 (2.1%) | 19 (1.5%) |
| Unknown | -- | -- | -- |
| Thyroid medications, n (%) | -- | -- | -- |
| Beta-blockers, n (%) | 473 (17%) | 233 (15%) | 240 (19%) |
| Unknown | -- | -- | -- |
| Other anti-hypertensive medications, n (%) | -- | -- | -- |
| Unknown | -- | -- | -- |

Abbreviations: BMI = body mass index.
<sup>a</sup>Characteristics without an “Unknown” category had no missing data.
<sup>b</sup>-- indicates cells n ≤ 10 masked for confidentiality.

**Table 2.**
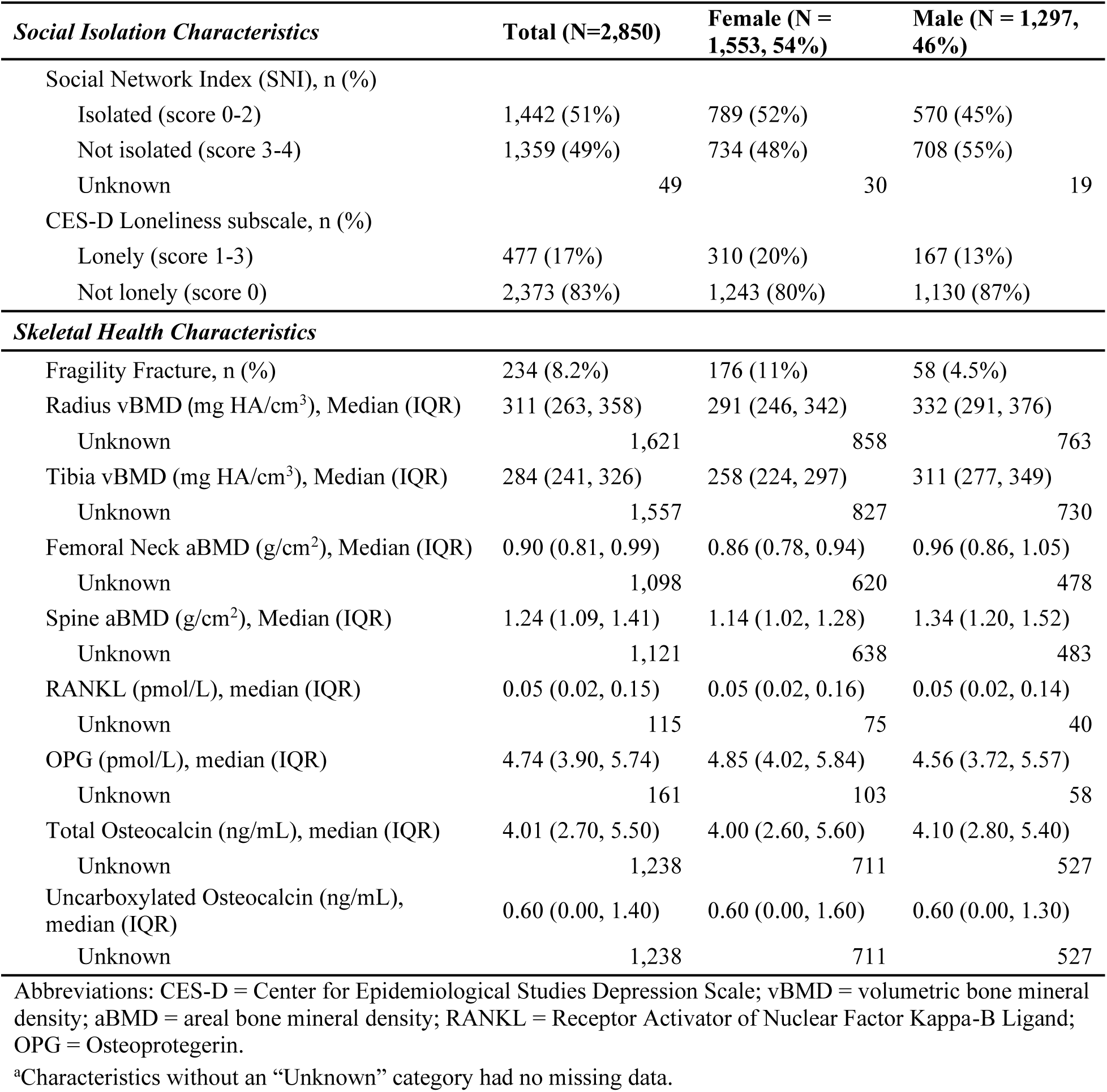
Social isolation and skeletal health characteristics of the FHS analytic sample^a^.

| ***Social Isolation Characteristics*** | **Total (N=2,850)** | **Female (N = 1,553, 54%)** | **Male (N = 1,297, 46%)** |
|---|---|---|---|
| Social Network Index (SNI), n (%) |  |  |  |
| Isolated (score 0-2) | 1,442 (51%) | 789 (52%) | 570 (45%) |
| Not isolated (score 3-4) | 1,359 (49%) | 734 (48%) | 708 (55%) |
| Unknown | 49 | 30 | 19 |
| CES-D Loneliness subscale, n (%) |  |  |  |
| Lonely (score 1-3) | 477 (17%) | 310 (20%) | 167 (13%) |
| Not lonely (score 0) | 2,373 (83%) | 1,243 (80%) | 1,130 (87%) |
| ***Skeletal Health Characteristics*** |  |  |  |
| Fragility Fracture, n (%) | 234 (8.2%) | 176 (11%) | 58 (4.5%) |
| Radius vBMD (mg HA/cm<sup>3</sup>), Median (IQR) | 311 (263, 358) | 291 (246, 342) | 332 (291, 376) |
| Unknown | 1,621 | 858 | 763 |
| Tibia vBMD (mg HA/cm<sup>3</sup>), Median (IQR) | 284 (241, 326) | 258 (224, 297) | 311 (277, 349) |
| Unknown | 1,557 | 827 | 730 |
| Femoral Neck aBMD (g/cm<sup>2</sup>), Median (IQR) | 0.90 (0.81, 0.99) | 0.86 (0.78, 0.94) | 0.96 (0.86, 1.05) |
| Unknown | 1,098 | 620 | 478 |
| Spine aBMD (g/cm<sup>2</sup>), Median (IQR) | 1.24 (1.09, 1.41) | 1.14 (1.02, 1.28) | 1.34 (1.20, 1.52) |
| Unknown | 1,121 | 638 | 483 |
| RANKL (pmol/L), median (IQR) | 0.05 (0.02, 0.15) | 0.05 (0.02, 0.16) | 0.05 (0.02, 0.14) |
| Unknown | 115 | 75 | 40 |
| OPG (pmol/L), median (IQR) | 4.74 (3.90, 5.74) | 4.85 (4.02, 5.84) | 4.56 (3.72, 5.57) |
| Unknown | 161 | 103 | 58 |
| Total Osteocalcin (ng/mL), median (IQR) | 4.01 (2.70, 5.50) | 4.00 (2.60, 5.60) | 4.10 (2.80, 5.40) |
| Unknown | 1,238 | 711 | 527 |
| Uncarboxylated Osteocalcin (ng/mL), median (IQR) | 0.60 (0.00, 1.40) | 0.60 (0.00, 1.60) | 0.60 (0.00, 1.30) |
| Unknown | 1,238 | 711 | 527 |

Abbreviations: CES-D = Center for Epidemiological Studies Depression Scale; vBMD = volumetric bone mineral density; aBMD = areal bone mineral density; RANKL = Receptor Activator of Nuclear Factor Kappa-B Ligand; OPG = Osteoprotegerin.
<sup>a</sup>Characteristics without an “Unknown” category had no missing data.

### Associations Between Social Isolation and Bone Outcomes

There was no association between SI or loneliness and fracture risk in women (**Figure 1A**, **Table 3**), nor were there associations between either objective SI or loneliness and BMD in women (**Figure 1B**, **Table 3**). However, in men, loneliness was associated with fracture risk, more than doubling the risk for fracture (OR = 2.46; 95% CI, 1.25 to 4.57; *P* = 0.011) (**Figure 1A**, **Table 3**). Additionally, loneliness was associated with a decrease in male tibia vBMD (β = -14.13, 95% CI, -28.03 to -0.23; *P =* 0.046) (**Figure 1B**, **Table 3**). In our sensitivity model (Model 2), the association between loneliness and male fracture risk remained positive after adjusting for all potential covariates, although it dropped below the significance threshold (OR = 2.00; 95% CI, 0.95 to 3.90; *P* = 0.066) (**eTable 1**).

**Figure 1.**
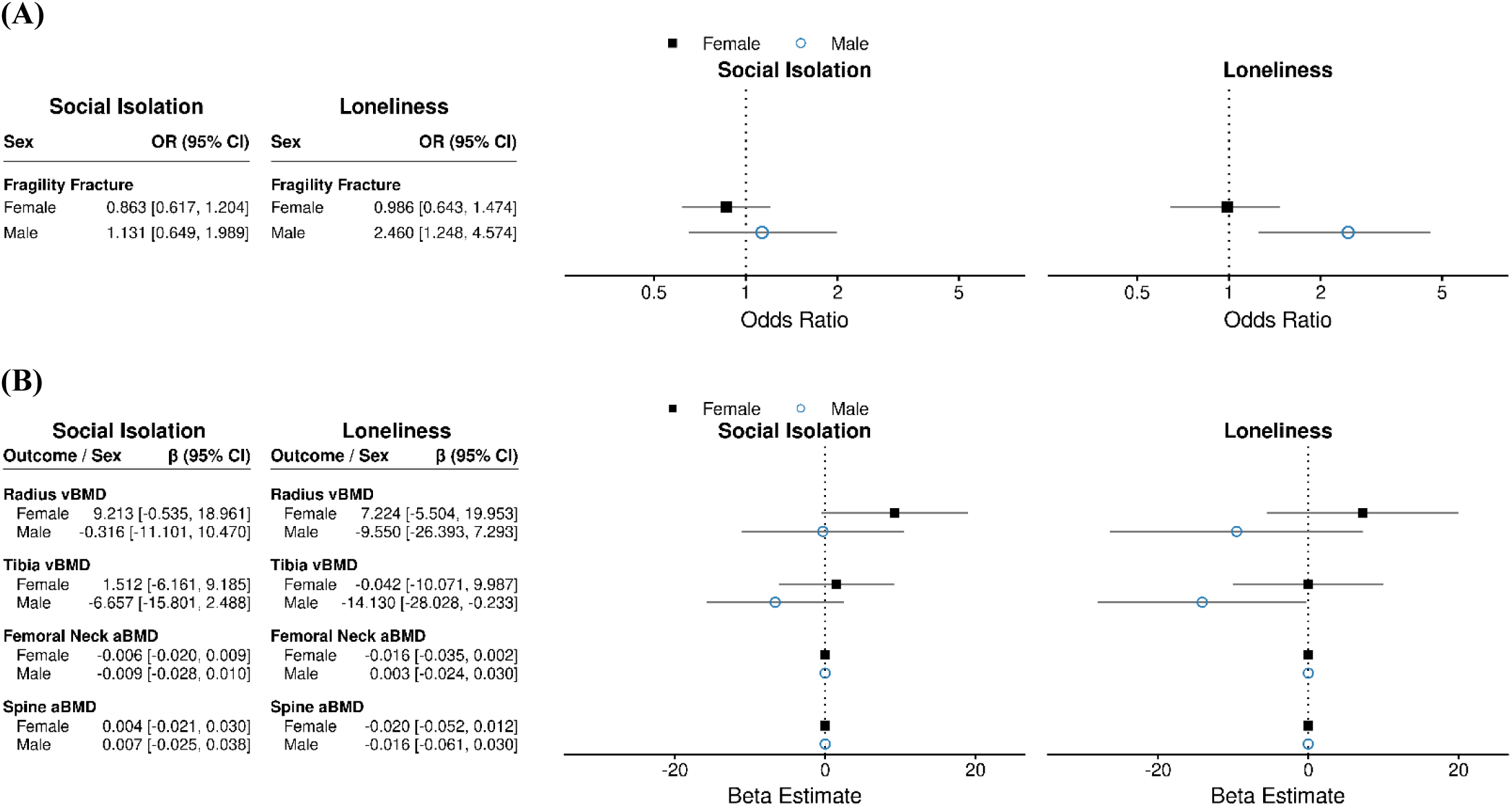
Forest Plots of Associations Between Objective and Subjective Social Isolation and Fragility Fracture and Bone Mineral Density. Results adjusted for age and body mass index, stratified by sex. Fragility fracture outcomes (**A**) modeled with Firth penalized logistic regression. BMD outcomes (**B**) modeled with linear regression. Error bars indicate 95% confidence intervals.

**Table 3.** Associations between social isolation factors and fracture, BMD, and bone turnover markers^a^.

|  | Model 0 - Unadjusted |  |  |  |  |  | Model 1 - Adjusted for Age and BMI |  |  |  |  |  |
|---|---|---|---|---|---|---|---|---|---|---|---|---|
|  | Females |  |  | Males |  |  | Females |  |  | Males |  |  |
|  | N | Estimate (95% CI) | p-value | N | Estimate (95% CI) | p-value | N | Estimate (95% CI) | p-value | N | Estimate (95% CI) | p-value |
| **Fragility Fracture** |  |  |  |  |  |  |  |  |  |  |  |  |
| Social Isolation | 1478 | 0.76 (0.54, 1.04) | 0.090 | 1260 | 0.97 (0.56, 1.69) | 0.918 | 1478 | 0.86 (0.62, 1.20) | 0.388 | 1260 | 1.13 (0.65, 1.99) | 0.655 |
| Loneliness | 1507 | 0.99 (0.65, 1.46) | 0.953 | 1279 | 2.29 (1.17, 4.21) | 0.017 | 1507 | 0.99 (0.64, 1.47) | 0.947 | 1279 | 2.46 (1.25, 4.57) | 0.011 |
| **Radius vBMD (mg HA/cm<sup>3</sup>)** |  |  |  |  |  |  |  |  |  |  |  |  |
| Social Isolation | 686 | 14.88 (4.62, 25.13) | 0.005 | 525 | 3.57 (-7.18, 14.33) | 0.514 | 686 | 9.21 (-0.53, 18.96) | 0.064 | 525 | -0.32 (-11.10, 10.47) | 0.954 |
| Loneliness | 693 | 12.18 (-1.34, 25.70) | 0.077 | 532 | -5.64 (-22.67, 11.38) | 0.515 | 693 | 7.22 (-5.50, 19.95) | 0.266 | 532 | -9.55 (-26.39, 7.29) | 0.266 |
| **Tibia vBMD (mg HA/cm<sup>3</sup>)** |  |  |  |  |  |  |  |  |  |  |  |  |
| Social Isolation | 716 | 4.59 (-3.70, 12.88) | 0.277 | 559 | -3.45 (-12.64, 5.75) | 0.462 | 716 | 1.51 (-6.16, 9.19) | 0.699 | 559 | -6.66 (-15.80, 2.49) | 0.153 |
| Loneliness | 724 | 3.28 (-7.59, 14.16) | 0.554 | 566 | -11.49 (-25.63, 2.65) | 0.111 | 724 | -0.04 (-10.07, 9.99) | 0.993 | 566 | -14.13 (-28.03, -0.23) | 0.046 |
| **Femoral neck aBMD (g/cm<sup>2</sup>)** |  |  |  |  |  |  |  |  |  |  |  |  |
| Social Isolation | 919 | 0.01 (-0.01, 0.03) | 0.259 | 808 | 0.00 (-0.02, 0.02) | 0.831 | 919 | -0.01 (-0.02, 0.01) | 0.438 | 808 | -0.01 (-0.03, 0.01) | 0.354 |
| Loneliness | 929 | -0.01 (-0.03, 0.01) | 0.281 | 816 | 0.01 (-0.02, 0.04) | 0.637 | 929 | -0.02 (-0.03, 0.00) | 0.089 | 816 | 0.00 (-0.02, 0.03) | 0.833 |
| **Spine aBMD (g/cm<sup>2</sup>)** |  |  |  |  |  |  |  |  |  |  |  |  |
| Social Isolation | 902 | 0.01 (-0.02, 0.03) | 0.552 | 804 | -0.01 (-0.04, 0.03) | 0.700 | 902 | 0.00 (-0.02, 0.03) | 0.745 | 804 | 0.01 (-0.02, 0.04) | 0.679 |
| Loneliness | 911 | -0.01 (-0.05, 0.02) | 0.425 | 811 | -0.03 (-0.07, 0.02) | 0.273 | 911 | -0.02 (-0.05, 0.01) | 0.223 | 811 | -0.02 (-0.06, 0.03) | 0.494 |
| **RANKL (pmol/L)** |  |  |  |  |  |  |  |  |  |  |  |  |
| Social Isolation | 1445 | 0.00 (-0.02, 0.03) | 0.738 | 1237 | 0.01 (-0.02, 0.03) | 0.456 | 1445 | 0.00 (-0.02, 0.02) | 0.991 | 1237 | 0.00 (-0.02, 0.03) | 0.713 |
| Loneliness | 1474 | 0.00 (-0.03, 0.03) | 0.763 | 1256 | 0.00 (-0.03, 0.04) | 0.838 | 1474 | 0.01 (-0.02, 0.03) | 0.729 | 1256 | 0.00 (-0.04, 0.04) | 0.966 |
| **OPG (pmol/L)** |  |  |  |  |  |  |  |  |  |  |  |  |
| Social Isolation | 1404 | -0.02 (-0.20, 0.16) | 0.803 | 1216 | -0.20 (-0.39, -0.00) | 0.049 | 1404 | 0.18 (0.01, 0.34) | 0.035 | 1216 | 0.05 (-0.12, 0.22) | 0.543 |
| Loneliness | 1431 | 0.15 (-0.08, 0.38) | 0.207 | 1233 | -0.01 (-0.30, 0.28) | 0.935 | 1431 | 0.17 (-0.03, 0.38) | 0.101 | 1233 | 0.14 (-0.11, 0.39) | 0.267 |
| **Total Osteocalcin (ng/mL)** |  |  |  |  |  |  |  |  |  |  |  |  |
| Social Isolation | 823 | -0.21 (-0.56, 0.14) | 0.233 | 755 | -0.39 (-0.71, -0.07) | 0.017 | 823 | -0.13 (-0.47, 0.22) | 0.479 | 755 | -0.43 (-0.75, -0.10) | 0.010 |
| Loneliness | 834 | 0.06 (-0.38, 0.51) | 0.778 | 764 | -0.25 (-0.71, 0.22) | 0.301 | 834 | 0.15 (-0.28, 0.59) | 0.489 | 764 | -0.26 (-0.73, 0.20) | 0.269 |
| **Uncarboxylated Osteocalcin (ng/mL)** |  |  |  |  |  |  |  |  |  |  |  |  |
| Social Isolation | 823 | -0.18 (-0.39, 0.02) | 0.081 | 755 | -0.21 (-0.39, -0.03) | 0.022 | 823 | -0.13 (-0.34, 0.07) | 0.199 | 755 | -0.21 (-0.39, -0.02) | 0.028 |
| Loneliness | 834 | -0.02 (-0.29, 0.24) | 0.855 | 764 | -0.19 (-0.45, 0.07) | 0.158 | 834 | 0.01 (-0.25, 0.27) | 0.947 | 764 | -0.18 (-0.44, 0.08) | 0.169 |

Abbreviations: vBMD = volumetric bone mineral density; aBMD = areal bone mineral density; RANKL = Receptor Activator of Nuclear Factor Kappa-B Ligand; OPG = Osteoprotegerin.
<sup>a</sup>Exposures measured at Exam 7. Fragility fractures measured from Exam 7-9. Radius and Tibia vBMD measured at Exam 9. Femoral neck and spine aBMD measured at Exam 8.

To assess how SI may impact bone formation and resorption, we investigated changes in markers of bone turnover. In women, objective SI was associated with an increase in OPG, a marker of bone formation (β = 0.18; 95% CI, 0.01 to 0.34; *P =* 0.035) (**Figure 2**, **Table 3**). Conversely, objective SI was associated with a decrease in total (β = -0.43; 95% CI, -0.75 to -0.10; *P =* 0.01) and uncarboxylated osteocalcin (β = -0.21; 95% CI, -0.39 to -0.02; *P =* 0.028) in men, also markers of bone formation (**Figure 2**, **Table 3**). The negative association between objective SI and both osteocalcin measures remained significant in our Model 2 sensitivity analysis (total osteocalcin: β = -0.41; 95% CI, -0.75 to -0.08; *P* = 0.016; uncarboxylated osteocalcin: β = -0.23; 95% CI, -0.43 to -0.04; *P* = 0.016) (**eTable 1**).

**Figure 2.**
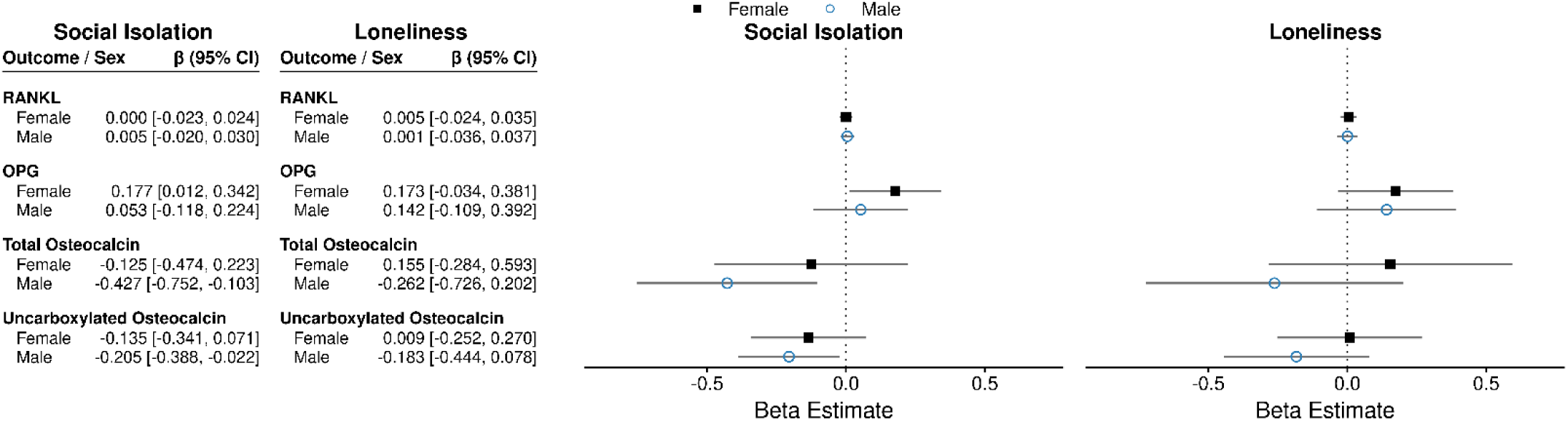
Forst Plot of Associations Between Objective and Subjective Social Isolation and Bone Turnover Markers. Results adjusted for age and body mass index, stratified by sex. Bone turnover outcomes modeled with linear regression. Error bars indicate 95% confidence intervals.

### Associations Between Social Isolation, Sex Hormones, and Inflammatory Markers

We examined the cross-sectional association between objective SI or loneliness and estradiol and testosterone in women, and testosterone in men, to understand if the sex-dependent effects of SI on bone are associated with changes in sex hormones (**eTable 2**). There was no significant association between objective SI or loneliness and estradiol, free testosterone, or total testosterone in either sex. Objectively isolated women had lower free testosterone levels, but the association was not significant (β = -0.067; 95% CI, -0.141 to 0.007; *P =* 0.075).

To determine if the effects of SI on bone could be a result of inflammatory changes, we tested the association between objective SI or loneliness and inflammatory markers. Objective SI was associated with higher 8-isoprostane in women (β = 0.177; 95% CI, 0.062 to 0.292; *P* = 0.003, *q* = 0.025) and higher interleukin 6 (IL-6) in men (β = 0.195; 95% CI, 0.085 to 0.304; *P =* 0.001, *q* = 0.005) (**eTable 3**). We tested if IL-6 mediated the significant associations between loneliness and bone outcomes in males. We found no significant mediating effect on fracture risk or vBMD (**eTable 4**).

## Discussion

Overall, the results of our study demonstrate that SI is associated with worsened skeletal health in a sex-specific manner. Loneliness was associated with increased fracture risk and lower vBMD in men at comparable levels to risk factors like smoking or consuming alcohol^42^. Objective SI was similarly associated with decreased bone formation markers in men. In contrast, SI had no effect on fracture risk or BMD in women but was associated with an increase in OPG. More women than men were isolated in our sample, indicating the sex differences in SI-induced bone loss are not a result of reduced exposure to SI in women. This finding is consistent with previous studies demonstrating that women are more likely to be lonely^29^, but SI’s effects on mortality risk^33^ and other physiological systems^31,32^ are worse in men.

There were no significant associations between SI and sex hormones in either sex. However, these analyses were cross-sectional rather than prospective, and estradiol measurements were unavailable for men, limiting our understanding of the role of sex hormones in SI-induced bone loss. Objective SI was, however, associated with increased inflammatory markers in both sexes, similar to previous studies^22^, but IL-6 did not mediate the association between loneliness and bone outcomes in men.

The results of the present study recapitulate our preclinical findings. We similarly found SI’s effects on bone microstructure and biomechanical properties in mice are male-specific^11,23^, and found no effect of SI on sex hormones^23^. We also found that SI increases bone formation markers in female mice^23^, similar to women in the present study. While female resilience to SI-induced bone loss observed in both human and rodent studies could be due to the general osteoprotective effects of estrogen^43^, over 95% of women in the present study were post-menopausal. Therefore, sex-specific factors beyond estrogen likely play an important role.

While loneliness and objective SI are associated with increased fracture risk and decreased bone formation markers respectively, we found few associations with BMD. Loneliness was associated with lower male tibia vBMD measured by pQCT, but there were no associations with aBMD measured by DXA. This is consistent with previous studies that found associations between SI and fracture or osteoporosis diagnosis, but not BMD^19,44^. While BMD is an important factor in fracture risk, individuals with normal BMD still suffer fragility fractures, often as a result of poor bone quality, including impaired material properties^45^. A lack of association with BMD may indicate that SI negatively impacts male bone quality rather than quantity. This is consistent with our preclinical findings that SI impairs material-level properties and energy-absorbing capabilities of bone in male mice^23^. This could suggest that DXA, the gold standard for osteoporosis screening and fracture prevention, may not capture individuals at risk for SI-induced bone loss and related fractures.

Interestingly, we observed differential effects of objective SI versus loneliness on bone parameters. While loneliness was associated with increased fracture risk and lower tibial BMD in men, objective SI was associated with increased and decreased markers of bone formation in women and men respectively. Similarly, objective SI was associated with increased inflammatory markers in both sexes.

Only two studies^20,22^ specifically considered differences in objective SI relative to loneliness in skeletal health, and found that both objective SI and loneliness increased osteoporosis^20,22^ and fracture risk^22^ to similar levels. The authors, however, did not look at BMD, bone turnover markers, or stratify analyses by sex. These data may indicate that lack of regular social connections and the subjective experience of loneliness act on different physiological pathways^46^ to impact bone health.

The association between objective SI and decreased osteocalcin in men is consistent with previous clinical and preclinical literature on other forms of stress. Stressful events are associated with decreased osteocalcin in humans^47^, and immobilization^48,49^ and chronic stress^49^ models decrease osteocalcin in rodents. These findings suggest SI may primarily impair bone by disrupting bone formation rather than by increasing resorption. Additionally, osteocalcin has an inverse relationship with glucocorticoids^50–52^. Endogenous glucocorticoids (i.e., cortisol) are integral to the stress response and typically increase with SI^53^, with known negative effects on bone^54^. Furthermore, the effects of glucocorticoids on bone are estrogen-dependent, and differ between sexes, potentially explaining the sex differences in SI-induced bone changes^43,55^, which should be investigated in future studies.

Objective SI was also associated with increased inflammatory markers, 8-isoprostane in women, and IL-6 in men. 8-isoprostane is a marker of oxidative stress^56^, and higher levels have been found in patients with major depressive disorder^57^. Additionally, sex differences in SI-induced cognitive deficits in rats have been linked to 8-isoprostane, with higher levels found in females^58^. Clinical studies have also found an association between SI and IL-6 levels^59^, including as a mediator between loneliness and gait speed^60^, and preclinical studies have implicated IL-6 in SI-induced brain deficits^61^. While our results found no significant mediation effect, our analysis was limited by the timing of variable collection, and could only assess IL-6, fracture, and vBMD in men, and should be replicated in future studies.

Our findings raise important clinical concerns. Older adult men may be more vulnerable to SI- induced bone loss but are also less likely to report feelings of isolation or loneliness^30^, and less likely to be screened for osteoporosis^62,63^. While women are more vulnerable to osteoporosis and fracture, approximately 1 in 5 men over the age of 50 will experience a fragility fracture^64^, and have higher mortality after fracture than women^63^. Yet screening guidelines for men are unclear^62^. The US Preventative Services Task Force recently concluded there was insufficient evidence to assess the benefits or harms of osteoporosis screening in men^63^. Furthermore, BMD measures via DXA may not capture the compromised bone properties that increase men’s SI-induced fracture risk. Future research should investigate how SI assessments could be incorporated to better predict fracture risk. Clinicians may also consider other screening tools when working with older adult men at risk for SI, such as blood-based bone turnover markers.

### Limitations

There were several limitations to this study. First, while we saw consistent associations between SI and bone outcomes in our primary Model 1, several of these associations did not retain significance in our sensitivity Model 2. Model 2 included a substantially larger covariate set, which yielded a smaller complete-case sample, likely reducing our statistical precision and attenuating some significant associations from Model 1. Future work should replicate our findings with larger sample sizes.

Additionally, while fractures and BMD were measured prospectively after SI exposures, most turnover markers and sex hormones were measured cross-sectionally at the same exam as SI measures. This makes it difficult to establish causality and the directionality of associations. Furthermore, our sample was 99% white with an average age of 60 years old, thus our results may not be applicable to more diverse demographic groups. Future studies should replicate our findings in diverse cohorts and examine how SI-induced bone loss and sex differences may change with age.

Collectively, the results of this study demonstrate that the association between SI and impaired skeletal health is sexually dimorphic, disproportionally affecting men. Loneliness was associated with increased fracture risk, and objective SI was associated with decreased markers of bone formation in older adult men but not women. Our findings underscore the need to consider SI as a major modifiable risk factor for male skeletal health in clinical practice.

## Supporting information

eMethods

eTables

## Data Availability

All FHS data was obtained via NIHLBI Biologic Specimen and Data Repository Information Coordinating Center (BioLINCC). The analytical code is available upon request to the authors.

## Acknowledgments

Author Contributions

SE contributed to the design, analysis, and interpretation of the data, and drafting of the manuscript. JD contributed to the analysis and interpretation of the data, and reviewing of the manuscript. PEM contributed to the design and analysis of the data, and the drafting of the manuscript. BMM, EB, and KS contributed to the interpretation of the data and drafting of the manuscript. CWL contributed to the design, acquisition, analysis, and interpretation of the data, and reviewing of the manuscript. RVM was responsible for the funding acquisition and oversight of the study, and the conception, design, acquisition, and interpretation of the data and the drafting of the manuscript.

## Data Availability

All FHS data was obtained via NIHLBI’s Biologic Specimen and Data Repository Information Coordinating Center (BioLINCC). This manuscript was not prepared in collaboration with investigators of the Framingham Heart Study and does not necessarily reflect the opinions or views of the Framingham Heart Study, Boston University, or NHLBI. The analytical code is available upon request.

## Other Contributions

The authors would like to thank Drs. Alexandra Hinton, PhD, MPH and Cara Frankenfeld, PhD from the MaineHealth Institute for Research for their advice and suggestions on the analysis of the FHS sample. The authors would also like to thank all of the participants and staff of the Framingham Heart Study for providing the data used in this study.

