## Supplementary material for "Social isolation as a sex-specific risk factor for skeletal health in the Framingham Heart Study": eMethods

### Social Isolation Measures

The Berkman–Syme Social Network Index (SNI) was used to evaluate objective social isolation. The SNI is a self-administered questionnaire that includes questions on number and frequency of social contacts. Responses to these questions are scored to calculate an index of social connectedness[1]. Scores ranged from 0 (most isolated) to 4 (most socially connected).

The CES-D “Loneliness” measure was used to evaluate subjective social isolation. The CES-D Loneliness measure is a self-administered questionnaire and asks about the participant’s feelings of loneliness in the last week scored from 0-3 with 0 = “rarely or none of the time” (less than 1 day/week) to 3 = “most or all of the time” (5-7 days/week).[2]

### Fragility Fracture Definition

Fragility fractures were defined as self-reported fractures excluding fingers, toes, skull, and face, pathological fractures, and those due to severe or high impact trauma.

### Sensitivity Analysis

A fully adjusted Model 2 was also evaluated as a sensitivity analysis. This model was adjusted for all likely social and physiological confounders available in the dataset with more than 10 individuals affected. These included age, body mass index (BMI), smoking status, education level, physical activity, antidepressant use, beta-blocker use, diabetes, cancer, and cardiovascular disease. Race/ethnicity and glucocorticoid use were available in the dataset but not included in the final Model 2 due to sparse data and model separation with limited stable estimation in the sex-stratified models. Model 2 analyses were conducted using complete-case analytical sample required for the complete covariate set. Additional fixed-sample sensitivity analyses were used to compare Model 0 and 1 estimates within the Model 2 complete-case analytical sample to assess whether differences between primary and fully adjusted estimates were attributable to covariate adjustment rather than analytical sample composition.

Age was reported as a continuous variable in years based on the patient date of birth, and BMI was also reported as a continuous variable calculated from the patient’s height and weight. Smoking status was self-reported as a binary yes/no variable. Education level was self-reported as a categorical variable of either “high school or less”, “some college or more”, or “unknown”. Physical activity was a continuous variable self-reported as the number of hours engaged in moderate activity per day. Medications and comorbidities were obtained from clinic data and were reported as categorical variables of either “no”, “yes”, or “unknown”. All potential confounders were measured or reported at Exam 7, apart from education level reported at Exam 8.

### Mediation Analysis

We tested if inflammatory markers significantly associated with loneliness from our regression models mediated significant associations between loneliness and bone outcomes via causal mediation analysis. Mediation analyses were limited to associations that were significant in the primary Model 1 and where the timing of the available variables supported the expected exposure to mediator to outcome order. Loneliness was measured at Exam 7 and inflammatory mediators were measured at Exam 8. Therefore, the final mediation analyses included male loneliness as the exposure, IL-6 inflammatory marker as the mediator, and fragility fracture occurring after Exam 8 and tibia vBMD measured at Exam 9 as the outcomes. Bone outcomes measured at or before Exam 8 were not included because the mediator was not measured before that outcome. For both fragility fracture and tibia vBMD, the mediator was modeled as a function of loneliness, age, and BMI. Tibia vBMD was modeled using linear regression, and fragility fracture was modeled using Firth logistic regression, with loneliness, the mediator, age, and BMI included as predictors. Average causal mediation effects (ACME), average direct effects (ADE), and total effects were estimated using nonparametric bootstrapping with 5,000 simulations. Analyses were conducted using complete cases for each outcome. Mediation analyses were performed using the mediation package in R.
